# Prevalence, Clinical Subtypes, and Associated Factors of Dry Eye Disease Among Pregnant Women in Dar es Salaam, Tanzania

**DOI:** 10.64898/2026.05.15.26353277

**Authors:** Gazilo Zacharia, Gasper Mung’ong’o, Flavian Shengeza, Nelson Swai, Heavenlight Masuki, Celina Mhina, Suzan Mosenene, Yusta Mtogo, Milka Mafwiri

## Abstract

**Background:** Dry eye disease (DED) is a multifactorial condition marked by tear film instability and ocular inflammation, causing symptoms like grittiness and blurred vision. The global prevalence of Dry eye disease among pregnant women ranges from 27.4% to 89.3% and in Africa it ranges between 20% and 50%. Hormonal changes, advanced maternal age, late pregnancy and prolonged screen time play an important part in its development.

**Methodology:** A hospital-based cross-sectional study among 380 pregnant women. Systematic sampling technique was used for recruitment at the antenatal clinic in Mnazi Mmoja Hospital in Dar es Salaam. Clinical dry eye tests were performed along with the administration of a symptom questionnaire that included demographic characteristics and the OSDI tool where OSDI ≥13 is the threshold for diagnosis of DED. Data were analyzed using Stata version 17.0 and Modified Poisson analyses identified factors associated with dry eye disease, with statistical significance set at p-value<0.05.

**Results:** A total of 380 pregnant women were recruited and analyzed with the mean age 31.7±6.7, 196 (51.6%) were aged 31-46 years. Most were married 273 (71.8%) and 211 (55.5%) had completed secondary education. Dry eye disease (DED) prevalence was 53.2% (48.8%-58.2%). Among those with DED (n=202), 112 (55%) had mild symptoms, 26 (13%) moderate, and 64 (32%) severe. The most common DED subtype was unclassified 72 (35.6%), followed by mixed (67, 33.2%), evaporative 50 (24.8%), and aqueous deficient 13 (6.4%). Significant associations with DED were: advanced gestation age (aPR=2.18 (1.550-3.072), p<0.001), being a housewife (aPR=1.48(1.179-1.857), p=0.001), use of visual display terminals (aPR=1.36(1.219-1.845), p=0.048), working in low humidity (aPR=2.62(1.698-4.045), p=0.001), and working in air-conditioned rooms (aPR=2.40(1.685-3.415), p=0.001). Secondary education was protective against DED (aPR = 0.668 (0.466-0.958), p = 0.028).

**Conclusion:** Approximately half of pregnant women have DED, with unclassified DED being the predominant subtype. Late gestation age, occupation, extended screen time, and working environment are significantly associated factors. It is essential to integrate DED screening into antenatal care and establish standardized protocols on DED management. Also, it is essential to promote lifestyle modifications such as reduction of screen time and avoiding dry environments.

## INTRODUCTION

Dry eye disease (DED) is a multifactorial ocular surface disorder defined by tear film homeostasis disruption (1). Patients experience discomfort, visual disturbance, and tear film instability that may damage the ocular surface (1). A vicious cycle of instability, hyperosmolarity, and inflammation perpetuates and worsens the condition (1). Global DED prevalence ranges from 5% to 50% due to methodological differences across studies (2). Sub-Saharan African populations show substantially higher burden estimates than global averages. A meta-analysis of African studies reported a pooled prevalence of 42.0% with regional variation (3).

Pregnancy involves profound physiological and hormonal shifts that have well-documented effects on ocular surface homeostasis and tear film stability (4,5). Fluctuations in estrogen, progesterone, and androgen levels during gestation alter both lacrimal gland secretory function and meibomian gland lipid production, creating conditions conducive to tear film disruption (4). Current evidence collectively suggests that globally DED affects 27.4% to 89.3% of pregnant women while in Africa it ranges between 20% and 50%, with the highest burden concentrated in the second and third trimesters (4,6).

The pathophysiology of DED in pregnancy reflects compounded hormonal changes on ocular surface glands. Elevated estrogen competitively binds androgen receptors, reducing testosterone’s trophic effect on meibomian gland function (7). Higher prolactin levels in the third trimester impair dihydrotestosterone-mediated sodium pump expression, reducing aqueous tear output (4,7). Pregnant women demonstrate greater meibomian gland loss than non-pregnant controls, supporting hormonal promotion of evaporative DED (8). These mechanisms produce four recognized clinical subtypes based on underlying gland dysfunction. Evaporative, aqueous deficient, mixed, and unclassified DED arise when mechanisms overlap or cannot be clearly delineated (6).

Diagnostic approaches in both clinical practice and research combine symptom scoring using the Ocular Surface Disease Index (OSDI) with objective homeostasis markers including tear breakup time (TBUT), Schirmer’s test, and standardised ocular surface staining to establish DED diagnosis and subtype classification (9,10). Composite biomarker approaches incorporating multiple objective parameters have been proposed to improve diagnostic precision beyond single-test assessments (11). Tear osmolarity measurement under standardised slit-lamp conditions adds further objectivity, though its measurement requires careful attention to examination-induced artefact (12).

There is a high burden of DED documented across African populations with an established physiological vulnerability of pregnant women to the condition. However, there is a lack of data on prevalence, risk factors, and clinical subtypes among pregnant women in Tanzania. This knowledge gap limits the development of context-appropriate early-detection and management protocols within Tanzanian antenatal care. The present cross-sectional study was therefore designed to estimate DED prevalence, associated factors, and clinical subtypes among pregnant women at Mnazi Mmoja Hospital. The findings are intended to inform improvements in maternal eye care in Tanzanian health facilities.

## METHODOLOGY

### Study Design

This was an analytical cross-sectional study conducted over a six-month period from July to December 2024 among 380 pregnant women attending the antenatal clinic (ANC) at Mnazi Mmoja Hospital, Dar es Salaam.

### Study Area

This study was conducted at Mnazi Mmoja Hospital in Dar es Salaam, which serves as a teaching facility for Muhimbili University of Health and Allied Sciences (MUHAS) and other universities in Tanzania. It is one of the largest health facilities in Ilala District, located at the city centre approximately 3.3 km from MUHAS. The facility has a well-organised antenatal clinic operating Monday to Friday each week, receiving approximately 350 to 400 pregnant women monthly for follow-up visits and 60 to 100 women for first visits; it serves both direct-attending clients and referrals from across Dar es Salaam. The hospital has a functional eye care unit equipped with slit-lamp biomicroscopes, ophthalmoscopes, and trial sets, staffed by optometrists and eye care nurses, making it suitable for systematic DED assessment.

### Sample Size Calculation

The minimum sample size was calculated using the Kish and Leslie formula for cross-sectional studies:

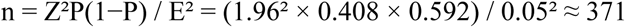

where n is the estimated minimum sample size, Z is the z-score at 95% confidence interval (1.96), P is the prior prevalence of DED in pregnancy (40.8%), derived from Asiedu et al. (2021) in a hospital-based ANC cohort in Ghana(6), and E is the margin of error (5%). The calculated minimum was 371 participants; the final recruitment target was set at 380 to account for potential non-response.

### Sampling Technique

Systematic random sampling was employed. The sampling frame of eligible women was constructed from the ANC register book (No. 6, MTUHA 6) after excluding those with systemic conditions. A total of 2,320 pregnant women attended the clinic during the study period. The sampling interval (K) was derived from the formula K = N/n, yielding K = 6; every sixth eligible participant was therefore selected. Data collection was carried out Monday to Friday of each study week, with approximately 10 to 15 participants selected per day.

### Study Population

The study population comprised all pregnant women attending Mnazi Mmoja ANC from July to December 2024.

### Eligibility Criteria

### Inclusion Criteria

1. All pregnant women aged 18 years and above attending Mnazi Mmoja ANC with a normal pregnancy during the study period.
2. Clinically confirmed pregnancy by urine pregnancy test and/or pelvic ultrasonography.

### Exclusion Criteria

1. Pregnant women with systemic conditions such as diabetes mellitus or systemic hypertension.
2. Pregnant women with ocular surface conditions, including blepharitis, allergic conjunctivitis, or pterygium greater than grade 2, which can affect tear film function.
3. Pregnant women on topical medications within three months prior to or at recruitment, including antihistamines or intraocular pressure-lowering agents.
4. Pregnant women with a history of ocular surgery, including glaucoma or corneal refractive procedures.
5. Pregnant women with eyelid abnormalities such as entropion, ectropion, or extensive ocular scarring.
6. Pregnant women who had worn contact lenses within four months prior to data collection.

### Data Collection

Data collection was conducted from 1 July to 31 December 2024. On each planned collection day at 10:00 hours, the register at the nursing station was reviewed to construct the daily sampling frame. Every sixth eligible participant was selected after on-site exclusion of women with systemic diseases, relevant systemic medications, or autoimmune conditions. Selected participants received a full explanation of the study and were enrolled following written informed consent. Priority was given to completing each participant’s routine ANC consultation before transferring to the eye clinic for ocular assessment.

Initial slit-lamp examination was performed to identify and exclude women with blepharitis, pterygium greater than grade II, eyelid abnormalities, or signs of allergic conjunctivitis. For enrolled participants, a standard structured interview-based questionnaire was administered to collect sociodemographic and obstetric data. Participants scoring 13 or more on the OSDI proceeded to full objective ocular assessment for DED signs.

### Clinical Assessment Procedures

#### Tear Film Breakup Time (TBUT)

TBUT was measured to quantify tear film stability. The test was performed at room temperature (24°C) with no fans or air conditioning. A fluorescein strip was moistened with normal saline, shaken to remove excess fluid, and applied by brief contact to the temporal one-third of the lower conjunctival sac of each eye (1,9,13). The stained cornea was viewed under slit-lamp cobalt blue illumination. The participant was instructed to blink twice, stare ahead, and blink once more; the interval from the final blink to the first appearance of a dry spot was measured with a stopwatch. Three readings were recorded and averaged; a mean TBUT below 10 seconds was considered positive for DED (6).

#### Ocular Surface Staining

Ocular surface staining assessed epithelial damage extent and severity. After TBUT measurement, fluorescein corneal staining from the same strip application was graded under cobalt blue illumination. Conjunctival staining was then performed using a lissamine green strip applied by brief contact to the temporal bulbar conjunctiva 3 mm from the temporal limbus (6,9). A one-to-two-minute interval was allowed before conjunctival grading under low-illumination white slit-lamp light. Both corneal and conjunctival staining were graded using the Oxford grading scale (9,10).

#### Schirmer’s Test 1

Schirmer’s test 1 was performed to measure basal and reflex tear secretion. Testing was at room temperature (24°C) with no fans or air conditioning. After mopping the eye dry, the folded end of a 35 mm by 5 mm pre-calibrated Whatman No. 41 filter paper strip was inserted without corneal contact into the junction between the lateral one-third and medial two-thirds of the lower fornix (1,9,13). Wetting extent after five minutes was recorded in millimetres; a reading below 15 mm was defined as positive for aqueous tear deficiency (9,10).

#### Meibomian Gland Assessment

Meibomian gland expressibility (MGE) and secretion quality (QMG) were assessed across eight central lower eyelid glands using tolerable digital pressure applied to the eyelid tarsus for 10 to 15 seconds (8,11). Examination was performed under slit-lamp diffuse illumination at 10x magnification. MGE was graded as: 0 = all glands producing lipid (normal); 1 = three to four glands producing; 2 = one to two glands producing; 3 = no glands producing. QMG was graded as: 0 = clear lipid easily expressed (normal); 1 = cloudy lipid (mild); 2 = cloudy lipid with particles (moderate); 3 = inspissated secretions (severe). The highest score across assessed glands was taken as the final score.

#### Diagnosis of DED and Clinical Subtypes

DED was diagnosed when OSDI ≥ 13 combined with at least one objective sign: TBUT below 10 seconds or Oxford-scale ocular surface staining above 3, consistent with criteria used in prior pregnancy-specific DED research in Africa (1,6). For subtype classification: Aqueous Deficient DED was assigned when TBUT below 10 seconds or staining above 3 was present alongside Schirmer’s 1 below 15 mm; Evaporative Dry Eye was diagnosed following positive MGD assessment; Mixed DED was assigned when both MGD and abnormal Schirmer’s 1 co-existed; Unclassified Dry Eye was assigned when Schirmer’s test exceeded 15 mm, TBUT was normal, and MGD was not evident (6,9). Objective assessments were performed on both eyes and findings from the worse eye were used for analysis.

### Statistical Analysis

Data were analysed using Stata version 17.0. Categorical variables were summarised using frequencies and proportions; continuous variables were summarised using means with standard deviations or medians as appropriate. Associations between DED and independent variables were first assessed using Chi-square or Fisher’s exact tests. Variables with p<0.2 in bivariable analysis were entered into a modified Poisson regression model with robust standard errors to estimate adjusted prevalence ratios (aPRs) and 95% confidence intervals. Statistical significance was set at p < 0.05. Prior to multivariable analysis, multicollinearity among exposure variables was assessed using pairwise correlations and variance inflation factors (VIF); no VIF exceeded 4, indicating no significant collinearity.

### Ethical clearance and consideration

The ethical approval to conduct this study was obtained from MUHAS Institutional Review Board on 22^nd^ April 2024 with clearance number MUHAS-REC-04-2024-2186. Permission to conduct this study was obtained from the regional administrative secretary then from the District Medical Officer (DMO) of Ilala and from the medical officer incharge (MOI) of Mnazi Mmoja Hospital. Written informed consent was obtained from all participants. Information obtained from patients was handled with confidentiality as no third party was allowed to have access to the information and during the interview participants were name-coded concealing their actual names. The patients were receiving standard and appropriate health care as per the hospital protocol. Participants were having the right to withdraw from the study at any time without any effect on their management plan.

## RESULTS

A total of 380 pregnant women were recruited and analyzed. The participants were aged between 18-46 years with the mean age (SD) of 31.7 ± (6.7) years. Slightly more than a half of the study participants 196 (51.6%) were aged 31-46 years. Majority were married 273 (71.8%) and 211 (55.5%) had completed secondary education. About 157 (41.3%) participants were office workers, as shown in Table 1.

**Table 1:**
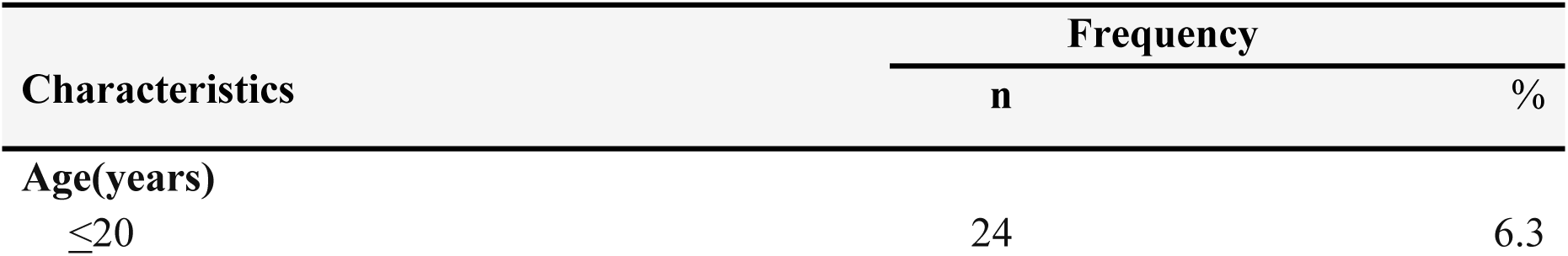
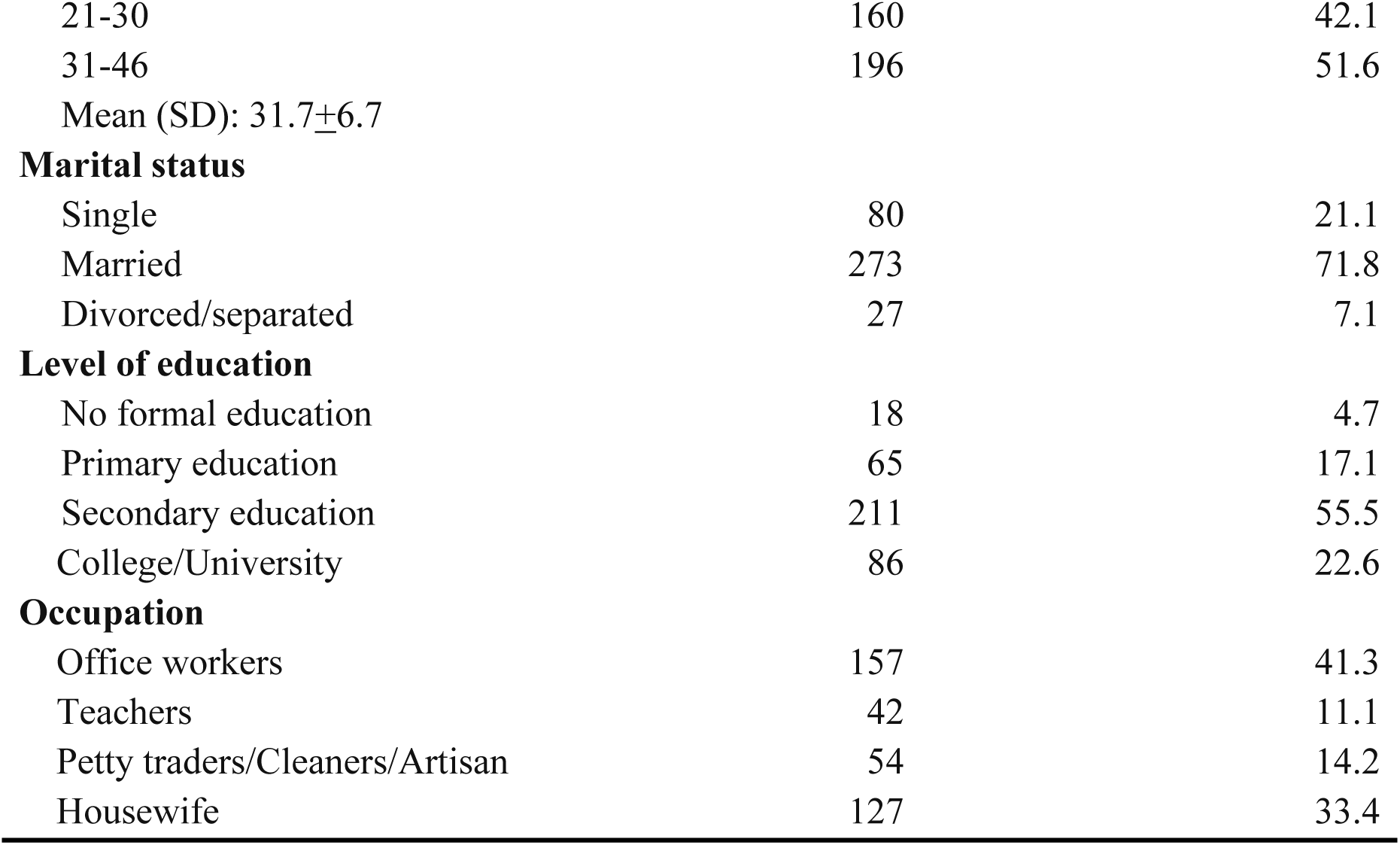
Social demographic characteristic of the study participants (N=380)

| Characteristics | Frequency |  |
| --- | --- | --- |
|  | n | % |
| Age(years) |  |  |
| ≤20 | 24 | 6.3 |
| 21-30 | 160 | 42.1 |
| 31-46 | 196 | 51.6 |
| Mean (SD): 31.7±6.7 |  |  |
| **Marital status** |  |  |
| Single | 80 | 21.1 |
| Married | 273 | 71.8 |
| Divorced/separated | 27 | 7.1 |
| **Level of education** |  |  |
| No formal education | 18 | 4.7 |
| Primary education | 65 | 17.1 |
| Secondary education | 211 | 55.5 |
| College/University | 86 | 22.6 |
| **Occupation** |  |  |
| Office workers | 157 | 41.3 |
| Teachers | 42 | 11.1 |
| Petty traders/Cleaners/Artisan | 54 | 14.2 |
| Housewife | 127 | 33.4 |

**Table 2:**
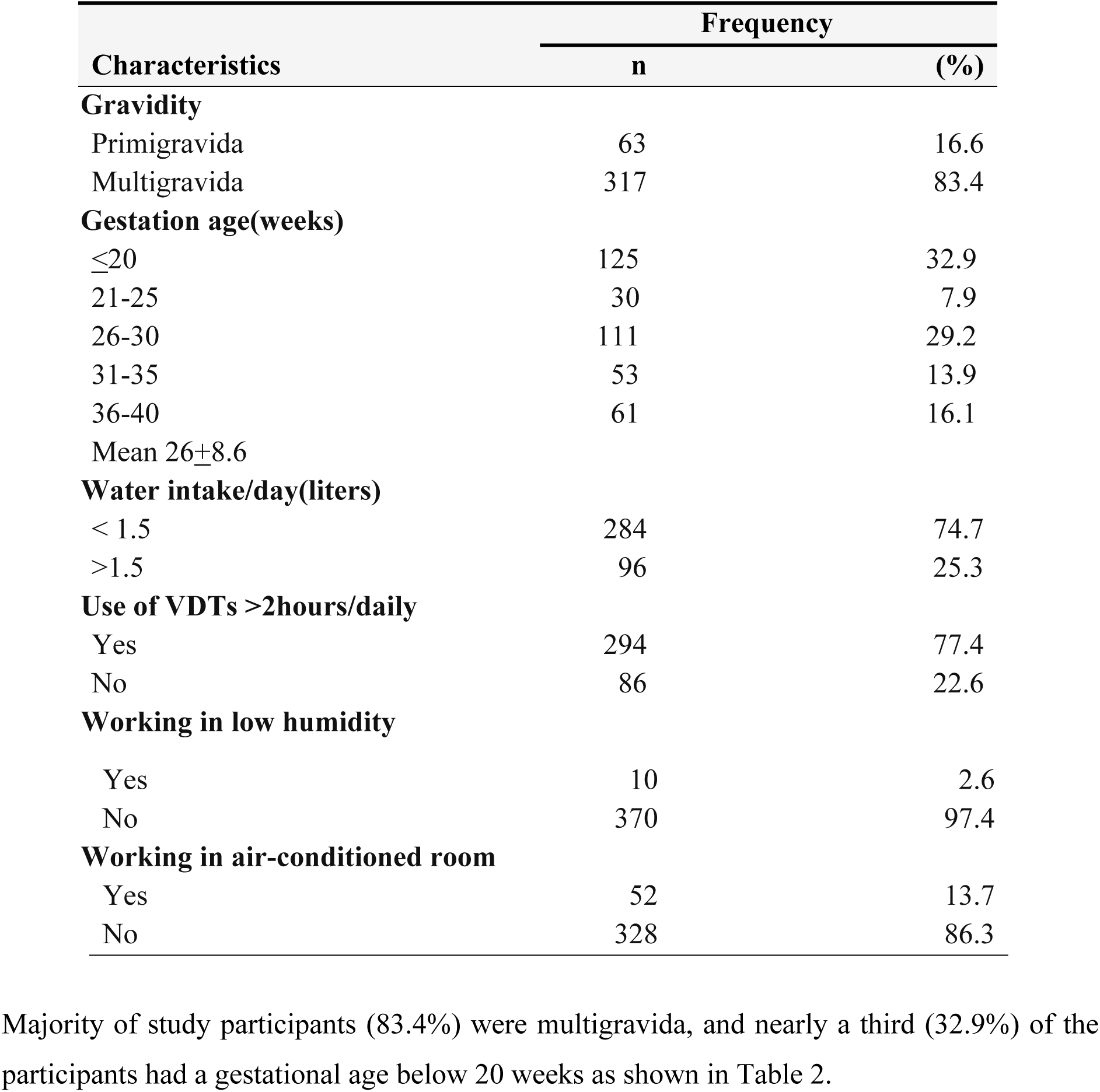
Distribution of baseline characteristics of the study participants (N=380)

| Characteristics | Frequency |  |
| --- | --- | --- |
|  | n | (%) |
| **Gravidity** |  |  |
| Primigravida | 63 | 16.6 |
| Multigravida | 317 | 83.4 |
| **Gestation age(weeks)** |  |  |
| ≤20 | 125 | 32.9 |
| 21-25 | 30 | 7.9 |
| 26-30 | 111 | 29.2 |
| 31-35 | 53 | 13.9 |
| 36-40 | 61 | 16.1 |
| Mean 26±8.6 |  |  |
| **Water intake/day(liters)** |  |  |
| < 1.5 | 284 | 74.7 |
| >1.5 | 96 | 25.3 |
| **Use of VDTs >2hours/daily** |  |  |
| Yes | 294 | 77.4 |
| No | 86 | 22.6 |
| **Working in low humidity** |  |  |
| Yes | 10 | 2.6 |
| No | 370 | 97.4 |
| **Working in air-conditioned room** |  |  |
| Yes | 52 | 13.7 |
| No | 328 | 86.3 |

Based on OSDI score, the proportion of pregnant women with DED was 53.2%, with a 95% confidence interval of (48.8%-58.2%).

More than half of the participants with DED 112(55%) experienced mild DED symptoms.

More than half of the participants with DED (59.9%) had reduced TBUT. (Figure 3)

**Figure 1:**
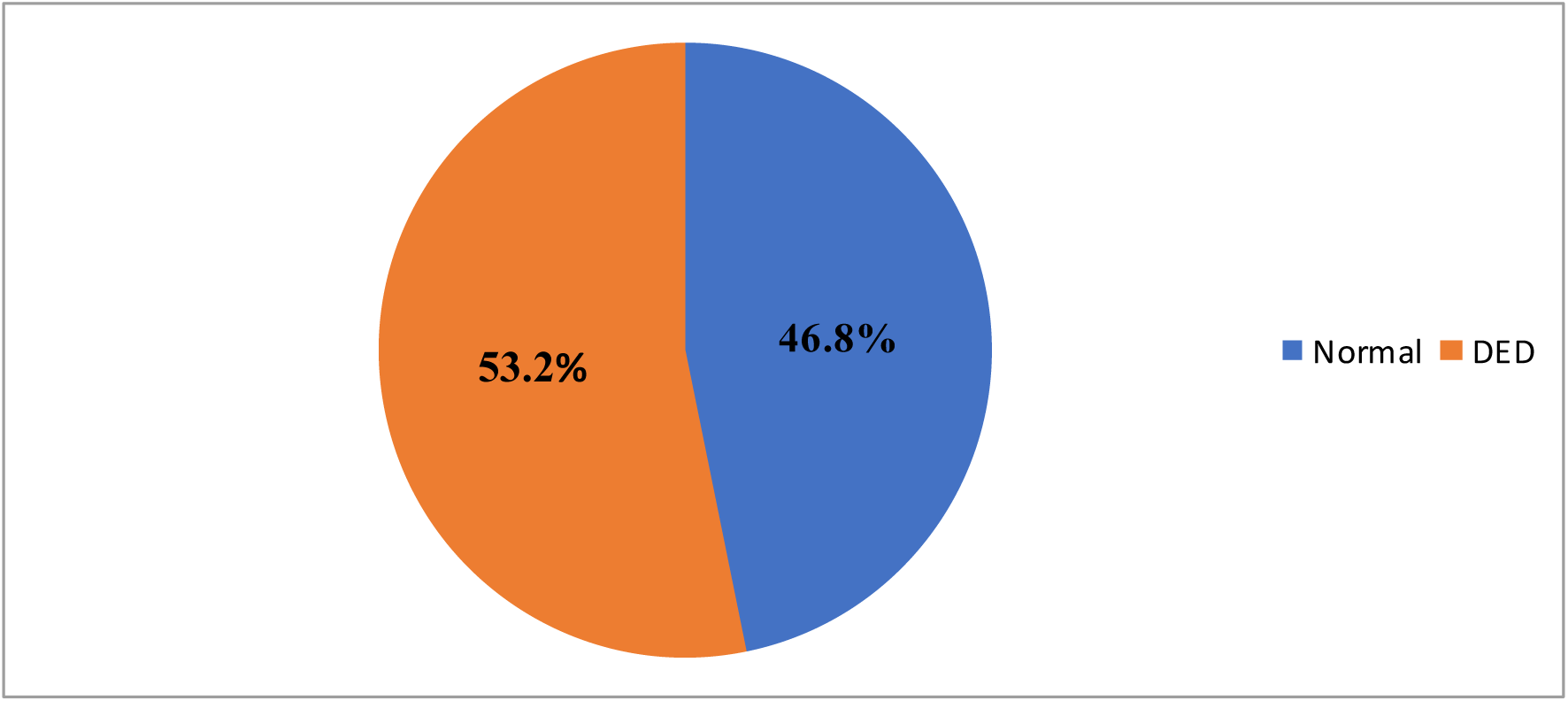
Proportion of pregnant women with DED based on the OSDI score.

**Figure 2:**
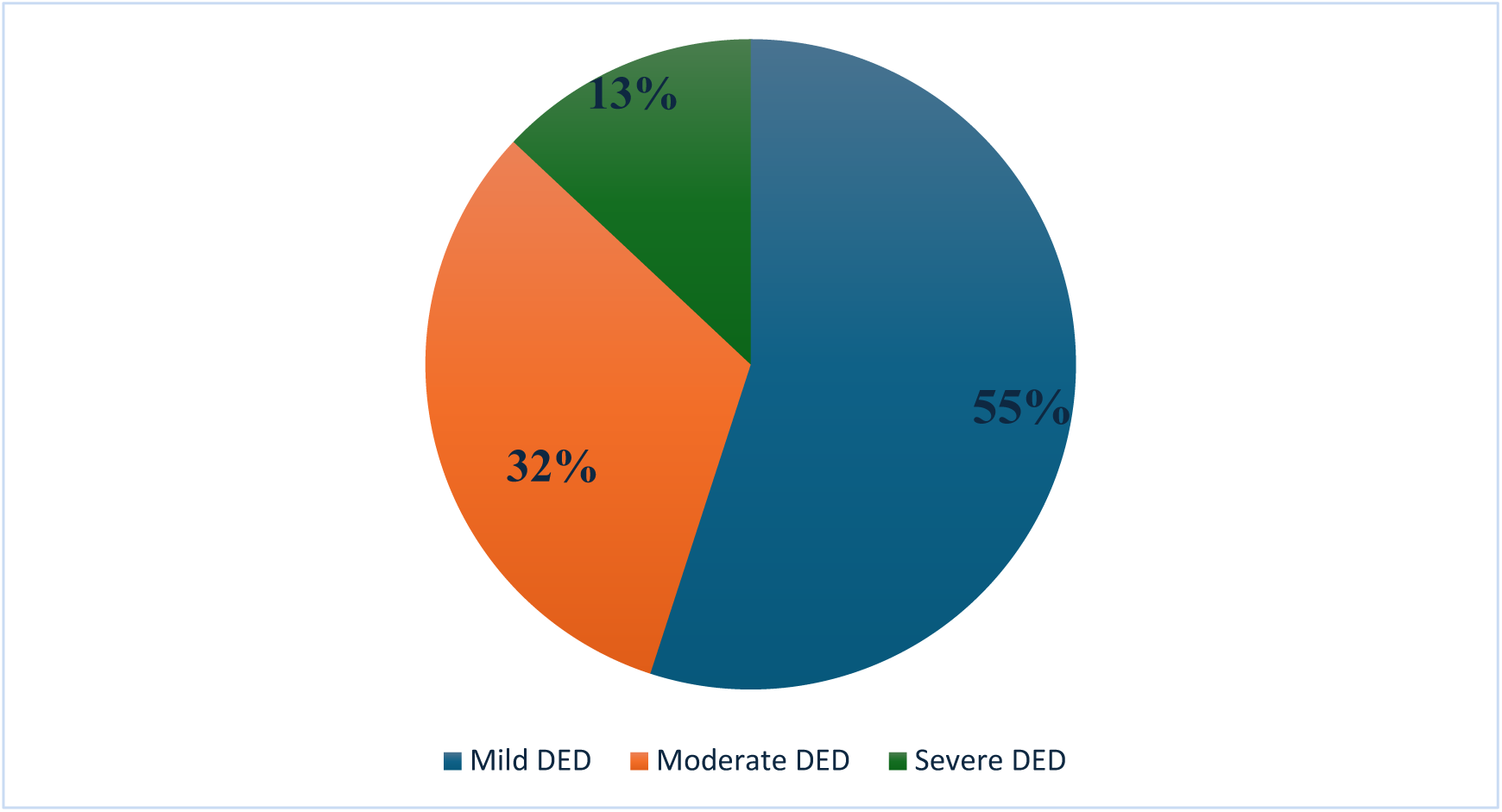
Proportion of pregnant women with DED according to the severity based on the OSDI score.

**Figure 3:**
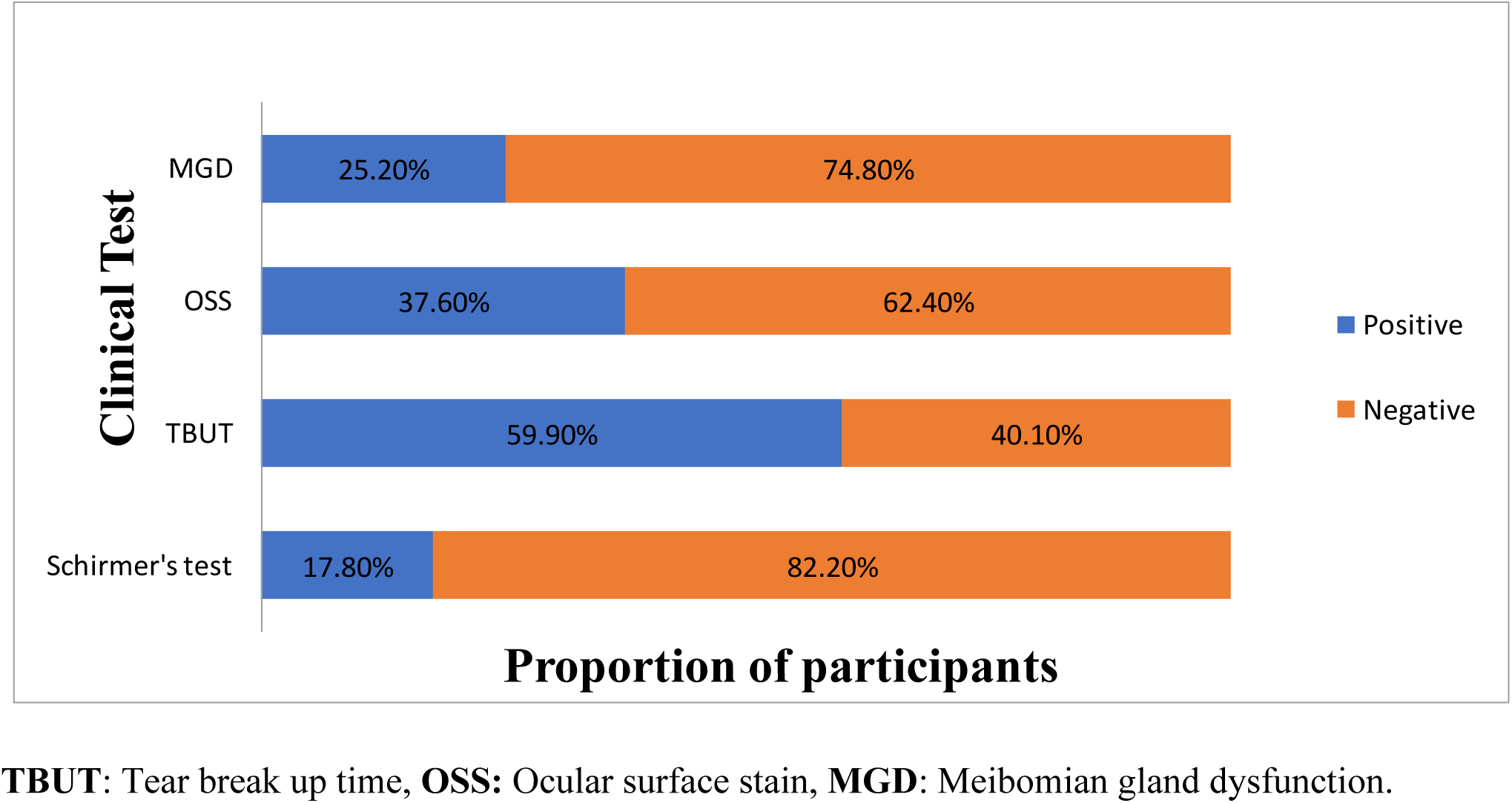
Proportion of pregnant women with DED according to the clinical test (N=202).

Unclassified DED was observed in 72(35.6%) participants, while aqueous deficiency dry eye was observed in 6.4% of the participants, as shown in Figure 4 below.

**Figure 4:**
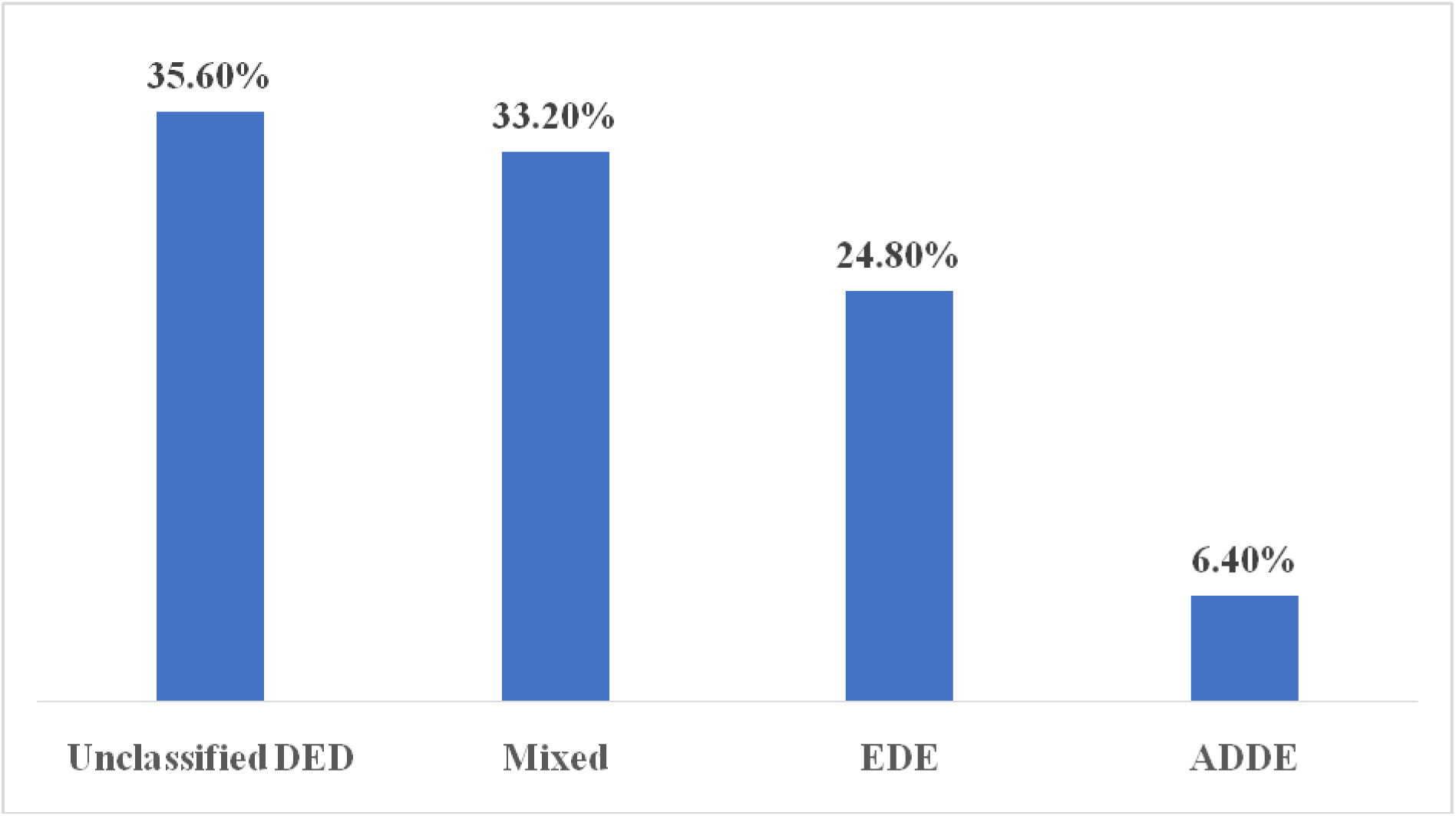
Proportion of clinical subtypes of DED (N=202)

In the multivariable analysis, being a housewife, increasing gestational age, use of visual display terminals (VDTs), and working in low-humidity and air-conditioned environments were independently associated with dry eye disease (DED). Pregnant women who used VDTs had 1.355 times the prevalence of DED compared to those who did not use these devices (aPR = 1.355; 95% CI: 1.219–1.845). Pregnant women with advanced gestational age (36–40 weeks) had 2.182 times the prevalence of DED compared to those with gestational age below 20 weeks (aPR = 2.182; 95% CI: 1.550–3.072). Housewives had 1.480 times the prevalence of DED compared to office workers (aPR = 1.480; 95% CI: 1.179–1.857). Similarly, participants working in low-humidity environments and those working in air-conditioned settings had 2.621 times and 2.399 times the prevalence of DED, respectively, compared to those not exposed to these conditions (Table 4).

**Table 3:** Relationship between participants’ characteristics and DED among pregnant women (N=380).

| Characteristic | DED STATUS (OSDI SCORE) |  | P-values (X<sup>2</sup>) |
| --- | --- | --- | --- |
|  | Normal (%) | DED (%) |  |
| **Age(years)** |  |  |  |
| <20 | 16(66.7) | 8(33.3) | 0.059 |
| 21-30 | 81(50.6) | 79(49.4) |  |
| 31-46 | 81(41.3) | 115(58.7) |  |
| **Marital status** |  |  |  |
| Single | 39(48.8) | 41(51.2) | 0.923 |
| Married | 126(46.2) | 147(53.8) |  |
| Divorced/separated | 13(48.1) | 14(51.9) |  |
| **Level of education** |  |  |  |
| No formal education | 6(33.3) | 12(66.7) | 0.001 |
| Primary education | 27(41.5) | 38(58.5) |  |
| Secondary education | 117(55.5) | 94(44.5) |  |
| College/University | 28(32.6) | 58(67.4) |  |
| **Occupation** |  |  |  |
| Office workers | 91(58.0) | 66(42.0) | 0.003 |
| Teachers | 14(33.3) | 28(66.7) |  |
| Petty traders/Cleaners/Artisan | 23(42.6) | 31(57.4) |  |
| Housewife | 50(39.4) | 77(60.6) |  |
| **Gravidity** |  |  |  |
| Primigravida | 36(57.1) | 27(42.9) | 0.097 |
| Multigravida | 142(44.8) | 175(55.2) |  |
| **Gestation age(weeks)** |  |  |  |
| < 20 | 93(74.4) | 32(25.6) | 0.001 |
| 21-25 | 6(20.0) | 24(80.0) |  |
| 26-30 | 49(44.1) | 62(55.9) |  |
| 31-35 | 7(13.2) | 46(86.8) |  |
| 36-40 | 23(37.7) | 38(62.3) |  |
| **Use of VDTs >2hours/daily** |  |  |  |
| No | 54(62.7) | 32(37.3) | 0.033 |
| Yes | 130(44.2) | 164(55.8) |  |
| **Water intake/day(liter)** |  |  |  |
| >1.5 | 50(52.1) | 46(47.9) | 0.234 |
| <1.5 | 128(45.1) | 156(54.9) |  |
| **Working in Low Humidity** |  |  |  |
| No | 177(47.8) | 193(52.2) | 0.022* |
| Yes | 1(10.0) | 9(90.0) |  |
| **Working in Air-conditioned room** |  |  |  |
| No | 170(51.8) | 158(48.2) | 0.001 |
| Yes | 8(15.4) | 44(84.6) |  |

*Note: Fisher's exact test \**

**Table 4:** Bivariable and multivariable analysis of the factors associated with DED.

| Variables | Bivariable Analysis |  | Multivariable Analysis |  |
| --- | --- | --- | --- | --- |
|  | cPR(95%CI) | P-value | aPR(95%CI) | P-value |
| **Maternal Age(years)** |  |  |  |  |
| <20 | (Ref) |  |  |  |
| 21-30 | 1.481(0.823-2.667) | 0.190 | 1.216(0.645-2.292) | 0.545 |
| 31-46 | 1.789(1.000-3.197) | 0.050 | 1.458(0.760-2.800) | 0.257 |
| **Marital status** |  |  |  |  |
| Married | Ref |  |  |  |
| Single | 0.952(0.748-1.211) | 0.687 |  |  |
| Divorced/separated | 0.963(0.658-1.408) | 0.846 |  |  |
| **Level of education** |  |  |  |  |
| No formal education | Ref |  |  |  |
| Primary education | 0.951(0.685-1.322) | 0.767 | 0.877 (0.596-1.290) | 0.505 |
| Secondary education | 0.730(0.513-1.038) | 0.080 | 0.668(0.466-0.958) | **0.028** |
| College/University | 0.977(0.614-1.556) | 0.923 | 1.012(0.707-1.448) | 0.950 |
| **Occupation** |  |  |  |  |
| Office workers | Ref |  |  |  |
| Teachers | 1.586(1.196-2.103) | 0.001 | 1.327(0.977-1.801) | 0.070 |
| Petty traders/Cleaners/Artisan | 1.366(0.617-1.833) | 0.078 | 1.230(0.911-1.924) | 0.093 |
| Housewife | 1.442(1.144-1.818) | 0.002 | 1.480(1.179-1.857) | **0.001** |
| **Gravidity group** |  |  |  |  |
| Primigravida | Ref |  |  |  |
| Multigravida | 1.288(0.952 -1.743) | 0.075 | 1.017(0.720-1.436) | 0.924 |
| **Gestation age in weeks** |  |  |  |  |
| ≤20 | Ref |  |  |  |
| 21-25 | 2.505(0.492-12.747) | 0.268 | 2.118(0.710-6.314) | 0.178 |
| 26-30 | 4.187(0.687-25.521) | 0.121 | 3.867(1.003-4.895) | **0.049** |
| 31-35 | 1.092(1.011-1.605) | 0.043 | 3.125(2.205-4.429) | **0.001** |
| 36-40 | 2.878(2.093-3.959) | 0.001 | 2.182(1.550-3.072) | **0.001** |
| **Use of VDTs >2hours/day** |  |  |  |  |
| No | Ref |  |  |  |
| Yes | 1.585(0.996- 2.103) | 0.052 | 1.355(1.219-1.845) | **0.048** |
| **Water (liter) /day** |  |  |  |  |
| > 1.5 | Ref |  |  |  |
| <1.5 | 1.146(0.869-2.199) | 0.353 |  |  |
| **Low Humidity** |  |  |  |  |
| No | Ref |  |  |  |
| Yes | 5.918(2.702-12.959) | 0.001 | 2.621(1.698-4.045) | **0.001** |
| **Air-conditioned room** |  |  |  |  |
| No | Ref |  |  |  |
| Yes | 1.121(1.073-2.169) | 0.046 | 2.399(1.685-3.415) | **0.001** |

Key: cPR; crude prevalence ratio, aPR; adjusted prevalence ratio, Ref; Reference category, CI;
Confidence Interval

## DISCUSSION

In this study, the proportion of pregnant women with dry eye disease (DED) was 53.2%. This estimate aligns closely with findings from a study conducted in India, where a prevalence of 48.5% was reported using a symptom-based approach, suggesting a substantial burden of DED among pregnant women in South Asian and comparable settings (14). Symptom-based tools such as the Ocular Surface Disease Index (OSDI), as used in our study, are known to yield higher prevalence estimates compared to sign-based approaches, which partly explains the relatively high burden observed across multiple countries (2,15).

However, the prevalence in our study was lower than that reported in a Nigerian study (78.4%), where participants were restricted to the third trimester and postpartum period (5). This contrast highlights the importance of gestational stage composition when comparing prevalence across settings. Studies conducted in late pregnancy or postpartum periods tend to report higher DED prevalence due to cumulative hormonal and physiological changes. In contrast, inclusion of women across all trimesters, as in our study, likely dilutes the overall prevalence because early pregnancy is associated with relatively preserved tear film function (16). Therefore, differences in sampling frames rather than true epidemiological variation may largely explain these discrepancies.

The study found unclassified DED to be the most common clinical subtype, accounting for 35.6% of cases. Similar patterns have been reported in studies conducted in the United States and Ghana, where a large proportion of patients could not be distinctly categorized into aqueous-deficient or evaporative subtypes (6,17). This reflects the complex and multifactorial nature of DED, particularly in pregnancy where hormonal, environmental, and behavioral factors interact. The Tear Film and Ocular Surface Society Dry Eye Workshop II (TFOS DEWS II) also recognizes that mixed or unclassified forms are common in clinical practice (1).

In addition, a greater proportion of participants exhibited features of evaporative dry eye associated with meibomian gland dysfunction (MGD) compared to aqueous-deficient dry eye. This pattern is consistent with findings from studies in Ghana and the United States, where evaporative mechanisms are frequently dominant, particularly in female populations (1,2,6). Environmental exposures such as low humidity and indoor air conditioning, which were also significant in our study, likely exacerbate tear film evaporation and contribute to this subtype distribution. Based on TFOS DEWS II recommendations, interventions such as lid hygiene, warm compresses, and moisture-retaining eyewear remain important and safe strategies for managing DED during pregnancy (1).

In terms of factors associated with DED, this study identified occupation (housewife), prolonged visual display terminal (VDT) use, low humidity, air-conditioned environments, and gestational age as significant predictors. These findings are broadly consistent with international evidence, though the magnitude of associations varies by setting.

Use of VDTs for more than two hours per day was significantly associated with DED (p = 0.048). Similar associations have been reported across multiple countries, including where prolonged screen exposure reduces blink rate and increases tear evaporation (18). While the direction of association is consistent globally, differences in digital device usage patterns and occupational demands may influence the strength of this relationship.

Being a housewife was strongly associated with DED (p < 0.001). This finding may reflect increased exposure to indoor environmental risk factors such as cooking smoke, poor ventilation, and prolonged indoor stays. Studies from Europe and other regions have similarly reported higher DED risk among individuals engaged in predominantly indoor activities, particularly when combined with screen exposure and limited air circulation (19). Additionally, employment status may influence access to healthcare services, potentially affecting early detection and management of DED (19).

Secondary education was found to be protective against DED (aPR = 0.668, p = 0.028). This may reflect higher health literacy and better adoption of protective behaviors such as reduced screen exposure, adequate hydration, and timely healthcare-seeking. Although the relationship between education and DED is not consistently reported across all settings, several studies suggest that improved health awareness contributes to reduced exposure to modifiable risk factors (20).

Environmental factors were also strongly associated with DED in this study. Participants exposed to low humidity and air-conditioned environments had significantly higher odds of DED (p < 0.001). These findings are consistent with studies conducted in Asia and other regions, where low ambient humidity accelerates tear evaporation and worsens meibomian gland dysfunction (18). In tropical urban environments such as Dar es Salaam, indoor climate control may create microenvironments that promote ocular surface dryness, even in otherwise humid climates.

Gestational age emerged as a significant predictor, with women in advanced stages of pregnancy being approximately three times more likely to have DED. This finding is consistent with studies from Ghana and India that report worsening tear film stability with advancing gestation (6,16). However, some studies using alternative tools such as the Standard Patient Evaluation of Eye Dryness (SPEED) questionnaire report weaker or inconsistent associations, highlighting the role of measurement differences in shaping observed outcomes (14). The use of OSDI in our study may have improved sensitivity for detecting mild symptoms common in pregnancy (15). Climatic differences between regions, such as those between Dar es Salaam and Chennai, may further contribute to variation in findings.

### Strengths and Weaknesses of the Study

This study employed a cross-sectional design, which limits causal inference between identified factors and DED. However, the use of both subjective (OSDI) and objective measures (TBUT, OSS, Schirmer’s test, and MGD assessment) strengthens the validity and reliability of findings and aligns with TFOS DEWS II recommendations for comprehensive DED assessment (1).

A further limitation is that objective clinical tests were only performed among participants with OSDI scores ≥13. This may have resulted in underestimation of prevalence by missing asymptomatic or sign-predominant cases, a limitation also noted in similar studies globally.

Environmental exposure assessment was another challenge, particularly in capturing individual-level humidity variations. This was partially mitigated by referencing regional data from the Tanzania Meteorological Authority. Additionally, the absence of biochemical and hormonal assays limited direct evaluation of hormonal influences; however, gestational age was used as a proxy, as supported in existing literature.

Despite these limitations, the study’s findings are consistent with global patterns of pregnancy-associated DED, reinforcing the role of hormonal, environmental, and behavioral factors. Differences across countries are likely attributable to variations in climate, study design, diagnostic tools, and gestational-stage sampling. These findings support the need for harmonized methodologies in future multi-country research to better understand and address DED in pregnancy.

## CONCLUSION

This study found a high burden of dry eye disease among pregnant women attending antenatal care in Dar es Salaam, with more than half of participants affected. The majority of cases were mild in severity, and unclassified DED was the predominant clinical subtype, reflecting the multifactorial and heterogeneous pathophysiology of ocular surface disease in pregnancy. Advancing gestational age, prolonged visual display terminal use, indoor environmental exposures including low humidity and air-conditioned settings, and housewife occupation were independently associated with increased DED risk. Secondary education was protective, likely through improved health literacy and adoption of preventive behaviors.

These findings position pregnancy as a period of heightened ocular surface vulnerability shaped by both hormonal physiology and modifiable environmental and behavioral factors. The high burden observed, particularly in the second and third trimesters, underscores the need to integrate eye health assessment into routine antenatal care in Tanzania and similar resource-limited settings.

## RECOMMENDATIONS

Routine DED symptom screening using the OSDI questionnaire should be incorporated into antenatal care visits, with particular attention to women in the second and third trimesters. Women identified as symptomatic should be referred for objective ocular surface assessment where capacity exists. Safe, pregnancy-appropriate management options including preservative-free artificial tears and lid hygiene practices should be made available within antenatal services.

Targeted counseling on DED risk factors and prevention should be delivered as part of antenatal health education. Key messages should address reducing prolonged screen time, practicing deliberate blinking during device use, maintaining adequate hydration, and improving indoor air quality through ventilation. Women who work in air-conditioned or low-humidity environments should receive specific guidance on protective measures such as humidifiers and regular screen breaks.

Basic eye care competencies should be integrated into training curricula for midwives and community health workers to enable early identification and appropriate referral of DED at the primary care level. Health facility managers should consider equipping antenatal clinics with validated symptom tools to support systematic screening without requiring specialist ophthalmic staff.

Longitudinal studies tracking tear film parameters across trimesters and into the postpartum period are needed to clarify the temporal relationship between gestational progression and DED development. Future research should incorporate hormonal profiling and objective environmental exposure measurements to better characterize the underlying mechanisms. Standardized diagnostic protocols combining symptom-based and objective measures will

improve comparability across settings and support the development of context-specific clinical guidelines for DED management in pregnancy.

## Authors’ contributions

· **Conceptualization:** Gazilo Zacharia, Milka Mafwiri, Yusta Mtogo
· **Methodology:** Milka Mafwiri, Suzan Mosenene, Yusta Mtogo, Gazilo Zacharia
· **Investigation (data collection):** Gazilo Zacharia
· **Formal analysis:** Gazilo Zacharia,
· **Writing – original draft:** Gazilo Zacharia, Gasper Mung’ong’o
· **Writing – review & editing:** Milka Mafwiri, Flavian Shengeza, Yusta Mtogo, Nelson Swai N, Celina Mhina, Suzan Mosenene, Heavenlight Masuki
· **Supervision:** Milka Mafwiri, Yusta Mtogo, Suzan Mosenene, Flavian Shengeza,
· **Project administration:** Gazilo Zacharia
· **Funding acquisition:** This research received no external funding

## Data Availability

All relevant data underlying the findings of this study are included within the manuscript and/or its supporting information files and are available from the corresponding author upon reasonable request.

## Acknowledgement

The authors express their sincere gratitude to all who contributed to the success of this study. We extend our appreciation to the Regional authorities of Dar Es Salaam, the District Medical Officer (DMO) of Ilala, and the Medical Officer In-Charge (MOI) of Mnazi Mmoja Hospital for their support and cooperation. We are deeply grateful to Ms. Salome Sallu (Head of RCH Unit), Mr. Japhet Magasa (Optometrist), and all staff at both the Eye Clinic and Antenatal Clinic at Mnazi Mmoja Hospital for their invaluable assistance during data collection. Finally, we are deeply grateful to all study participants for their invaluable contributions, which were essential to the completion of this research. This study was conducted as part of a Master’s of Medicine degree in Ophthalmology at Muhimbili University of Health and Allied Sciences.

## References

1. Craig JP, Nichols KK, Akpek EK, Caffery B, Dua HS, Joo CK, et al. TFOS DEWS II Definition and Classification Report. Ocul Surf. 2017;15(3):276–83.

2. Stapleton F, Alves M, Bunya VY, Jalbert I, Lekhanont K, Malet F, et al. TFOS DEWS II Epidemiology Report. Ocul Surf. 2017;15(3):334–65.

3. Akowuah PK, Kobia-Acquah E. Prevalence of dry eye disease in Africa: a systematic review and meta-analysis. Optom Vis Sci. 2020;97(12):1089–98.

4. Jaruchowska M, Przybek-Skrzypecka J, Skrzypecki J. Pregnancy and Dry Eye Syndrome: A Review for Clinical Practice. Int J Mol Sci. 2025;26(20):9990.

5. Nkiru Z N, Stella O, Udeh N, Polycarp U A, Daniel C N, Ifeoma R E. Dry eye disease: A longitudinal study among pregnant women in Enugu, south east, Nigeria. Ocul Surf. 2019;17(3):458–63.

6. Asiedu K, Kyei S, Adanusa M, Ephraim RKD, Animful S, Ali-Baya SK, et al. Dry eye, its clinical subtypes and associated factors in healthy pregnancy: A cross-sectional study. PLoS One. 2021 Oct;16(10).

7. Kyei S, Ephraim RKD, Animful S, Adanusa M, Ali-Baya SK, Akorsah B, et al. Impact of serum prolactin and testosterone levels on the clinical parameters of dry eye in pregnant women. J Ophthalmol. 2020;2020(1):1491602.

8. Kunduracı MS, Koçkar A, Helvacıoğlu Ç, Kırık F, Karakuş Hacıoğlu G, Akçay BİS. Evaluation of dry eye and meibomian gland function in pregnancy. Int Ophthalmol. 2023;43(11):4263–9.

9. Alves M, Reinach PS, Paula JS, Cruz AA V, Bachette LG, Faustino J, et al. Comparison of Diagnostic Tests in Distinct Well-Defined Conditions Related to Dry Eye Disease. PLoS One. 2014;9(5):e97921.

10. Szczęsna-Iskander DH, Muzyka−Woźniak M, Quintana CL. The Efficacy of Ocular Surface Assessment Approaches in Evaluating Dry Eye Treatment With Artificial Tears. Sci Rep. 2022;12(1).

11. Gala-Núñez C, Ortiz-Peregrina S, Castanera-Gratacós D, Anera RG. Development of a Dry Eye Index as a New Biomarker of Dry Eye Disease. Ophthalmic Physiol Opt. 2024;44(7):1472–83.

12. Song MS, Park J, Paik HJ, Kim DH. Effects of Slit Lamp Examination on Tear Osmolarity in Normal Controls and Dry Eye Patients. Bioengineering. 2025;12(10):1124.

13. Wolffsohn JS, Arita R, Chalmers R, Djalilian A, Dogru M, Dumbleton K, et al. TFOS DEWS II diagnostic methodology report. Ocul Surf. 2017;15(3):539–74.

14. Anantharaman D, Radhakrishnan A, Anantharaman V. Subjective Dry Eye Symptoms in Pregnant Women-A SPEED Survey. J Pregnancy. 2023;2023:17–21.

15. Schiffman RM, Christianson MD, Jacobsen G, Hirsch JD, Reis BL. Reliability and validity of the ocular surface disease index. Arch Ophthalmol. 2000;118(5):615–21.

16. Rizyal A, Shrestha B, Khadka A. Dry Eye Disease among Pregnant Women at a Tertiary Care Hospital in Kathmandu. Nepal Med Coll J. 2020 Nov;22(3):146–52.

17. Lemp MA, Crews LA, Bron AJ, Foulks GN, Sullivan BD. Distribution of aqueous-deficient and evaporative dry eye in a clinic-based patient cohort: A retrospective study. Cornea. 2012;31(5):472–8.

18. Al-Mohtaseb Z, Schachter S, Shen Lee B, Garlich J, Trattler W. The relationship between dry eye disease and digital screen use. Clin Ophthalmol. 2021;3811–20.

19. Bazeer S, Jansonius N, Snieder H, Hammond C, Vehof J. The relationship between occupation and dry eye. Ocul Surf. 2019;17(3):484–90.

20. McCann P, Abraham AG, Mukhopadhyay A, Panagiotopoulou K, Chen H, Rittiphairoj T, et al. Prevalence and Incidence of Dry Eye and Meibomian Gland Dysfunction in the United States. Jama Ophthalmol. 2022;140(12):1181.

